# 40 Hz Auditory Steady-State Responses Predict Clinical Outcomes in Clinical-High-Risk Participants: A MEG Study

**DOI:** 10.1101/2020.09.25.20201327

**Authors:** Tineke Grent-‘t-Jong, Ruchika Gajwani, Joachim Gross, Andrew I. Gumley, Rajeev Krishnadas, Stephen M. Lawrie, Matthias Schwannauer, Frauke Schultze-Lutter, Peter J. Uhlhaas

## Abstract

**Objective:** To examine whether 40-Hz Auditory Steady-State Responses (ASSRs) are impaired in participants at clinical high-risk for psychosis (CHR-P) and predict clinical outcomes.

**Method:** Magnetoencephalography data were collected during a 40-Hz ASSR paradigm for a group of 116 CHR-P participants, 33 first-episode-psychosis patients (FEP, 15 antipsychotic-naïve), a psychosis-risk-negative group (CHR-N: n=38) and 49 healthy controls. Analysis of group differences of 40-Hz Inter-trial-phase-coherence (ITPC) and 40-Hz amplitude focused on right Heschl’s gyrus [RHES], superior temporal gyrus, hippocampus [RHIP], and thalamus [RTHA], after establishing significant activations during 40-Hz stimulation. Linear regression and linear discriminant analyses (LDA) were used to predict clinical outcomes, including transition to psychosis and persistence of attenuated psychotic symptoms (APS) in CHR-Ps.

**Results:** CHR-P and FEP-patients were impaired in 40-Hz amplitude in RTHA and RHIP. In addition, FEP-patients were impaired in 40-Hz amplitude in RHES and CHR-Ps in 40-Hz ITPC in RHES. 40-Hz ASSR deficits were pronounced in CHR-Ps who later transitioned to psychosis (n=13) or showed persistent APS (n=40). Importantly, both APS-persistence and transition to psychosis were predicted by 40-Hz ASSR impairments. Classification accuracy was 73.7% for non-persistent-APS and 72.5% for persistent-APS group (Area under the curve (AUC)=0.842). For transition risk to psychosis, classification accuracy was 76.7% and 53.8% for non-transitioned and transitioned CHR-P participants, respectively (AUC=0.810).

**Conclusions:** Our data indicate that deficits in gamma entrainment in primary auditory cortex and subcortical areas constitute a potential biomarker for predicting clinical outcomes in CHR-P participants.

## Introduction

The brain’s endogenous rhythmic activity represents a fundamental feature of large-scale circuits that has been implicated in cognition and behavior as well as in psychiatric syndromes, such as schizophrenia (1). One way to probe neural oscillations is through entrainment by exogenous sources, such as visual or auditory stimuli or brain stimulation (2). Current theories suggest that exogenous entrainment interacts with ongoing oscillatory of neural circuits that could provide possible treatment targets for brain disorders (3).

The Auditory Steady-State Response (ASSR) is an evoked oscillation that is entrained to the frequency and phase of temporally modulated auditory stimuli (4). ASSRs typically show a peak frequency at around 40 Hz, suggesting an auditory “resonant” frequency induced by external periodic stimulation (5). Magnetoencephalography (MEG), Positron Emission Tomography, intracranial recordings and functional Magnetic Resonance Imaging (fMRI) studies located the generators of the 40-Hz ASSR in medial areas of the primary auditory cortex that are distinct from those underlying transient auditory components (6), as well as in hippocampus (7), thalamus and brainstem regions (8, 9).

One important application of the 40-Hz ASSR has been in schizophrenia because of the potential importance of gamma-band (30-200 Hz) oscillations in explaining cognitive deficits (10). Gamma-band oscillations have been hypothesized to establish communication between distributed neuronal ensembles (11) and their impairment to underlie the pronounced cognitive and perceptual dysfunctions in schizophrenia (12). This view is consistent with data indicating that rhythm-generating parvalbumin-positive (PV+) γ-aminobutyric acid (GABA) interneurons and N-methyl-d-aspartate (NMDA) receptors are impaired in schizophrenia (13), highlighting the potential of gamma-band oscillations to provide insights into circuit abnormalities.

Currently, there is robust evidence that both amplitude and phase of 40-Hz ASSRs are impaired in schizophrenia (14, 15). There is mixed evidence, however, for the presence of 40-Hz ASSRs deficits in both clinical high risk for psychosis (CHR-P) individuals (16-19) and patients with first-episode psychosis (FEP) (18, 20). In addition, it is also currently unclear whether 40-Hz ASSRs could constitute a possible biomarker for the early detection and diagnosis of emerging psychosis.

To address this fundamental question, we applied a state-of-the-art MEG approach to examine 40-Hz ASSRs in CHR-Ps as well as FEP-patients. MEG is characterized by an improved signal-to-noise ratio for measurements of high-frequency oscillations compared to electroencephalography (EEG) (21), and is ideally suited for source-reconstruction. Source-level estimation of ASSR impairments is essential as signal strength has been shown to be higher at source-than sensor-level (22, 23). Crucially, it allows the unmixing of signal contributions from different source-generators that are obscured at sensor-level by individual differences in source-orientation and field spread (24).

In the current study, we investigated MEG-recorded virtual-channel 40-Hz ASSRs in a group of 116 CHR-Ps as, 33 FEP patients as well as 38 participants with substance-abuse and affective disorders (CHR-N) and compared them to 49 healthy controls. We predicted that FEP and CHR-P participants would be characterized by a circumscribed dysfunction of 40-Hz ASSRs in auditory cortex and subcortical stimulus-entrained brain regions, which would be closely linked to clinical outcomes. Specifically, we hypothesized that impaired 40-Hz ASSRs would predict persistence of attenuated psychotic symptoms (APS) as well as transition to psychosis in CHR-P participants.

## Methods and Materials

### Participants

A total of 236 participants were recruited from the Youth Mental Health Risk and Resilience (YouR) Study (25) and divided into four groups: 1) 116 participants meeting CHR-P criteria, (2) 38 participants characterized by non-psychotic disorders, such as affective disorders (n=11), anxiety disorders (n=16), eating disorders (n=1), and/or substance abuse (n=10) (CHR-N), 3) 33 patients with FEP (15 antipsychotic-naïve) and, 4) 49 healthy control participants (HC) without an axis I diagnosis or family history of psychosis.

CHR-P status at baseline was established by ultra-high risk criteria according to the Comprehensive Assessment of At Risk Mental States (CAARMS) Interview (26) and the Cognitive Disturbances (COGDIS) and Cognitive-Perceptive (COPER) basic symptoms criteria according to the Schizophrenia Proneness Instrument, Adult version (SPI-A) (27). FEP patients were assessed with the Structured Clinical Interview for DSM-IV (SCID) (see Table 1) (28) and with the Positive and Negative Symptom Scale (PANSS) (29). For all groups except FEP-patients, cognition was assessed with the Brief Assessment of Cognition in Schizophrenia (BACS) (30).

**Table 1.**
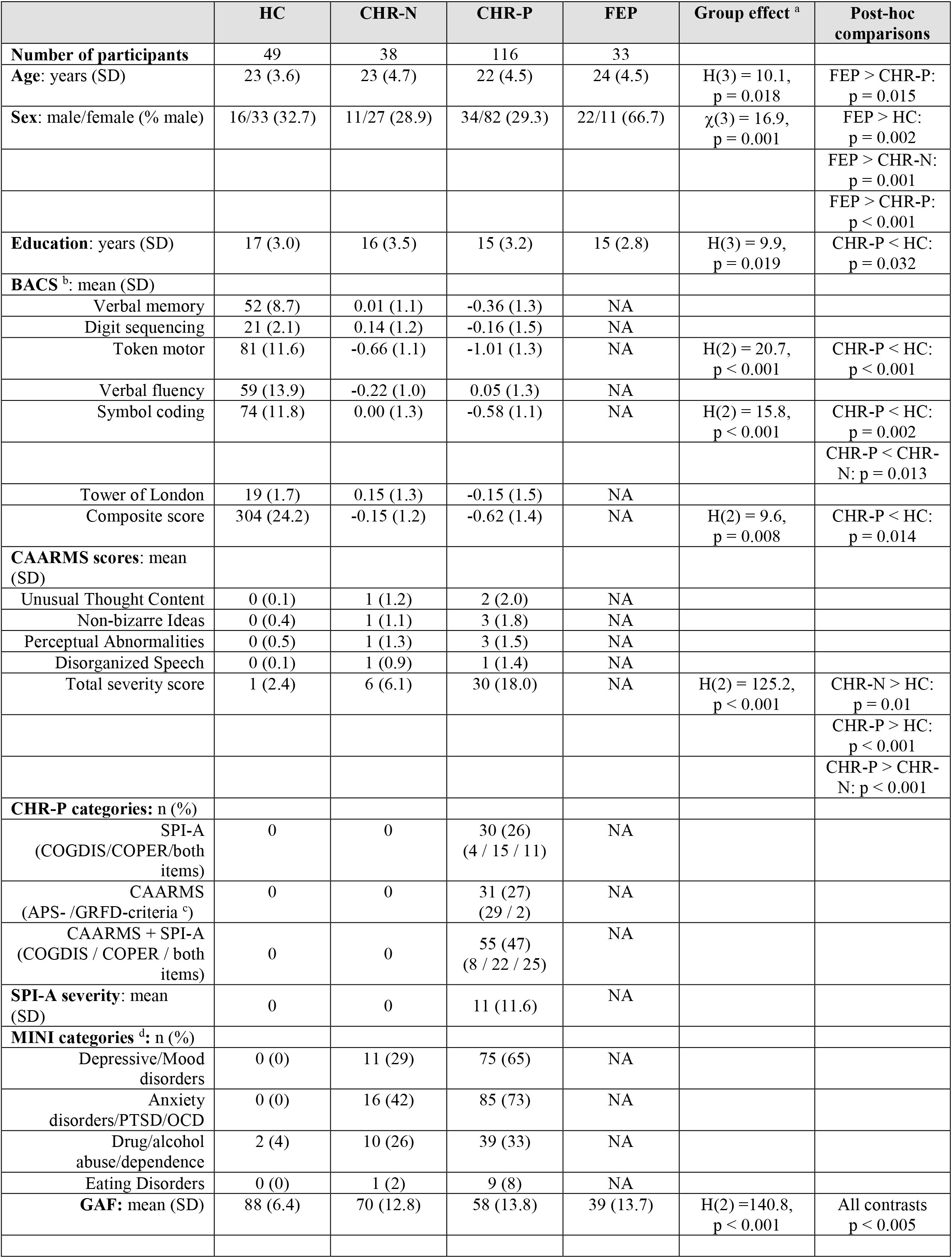
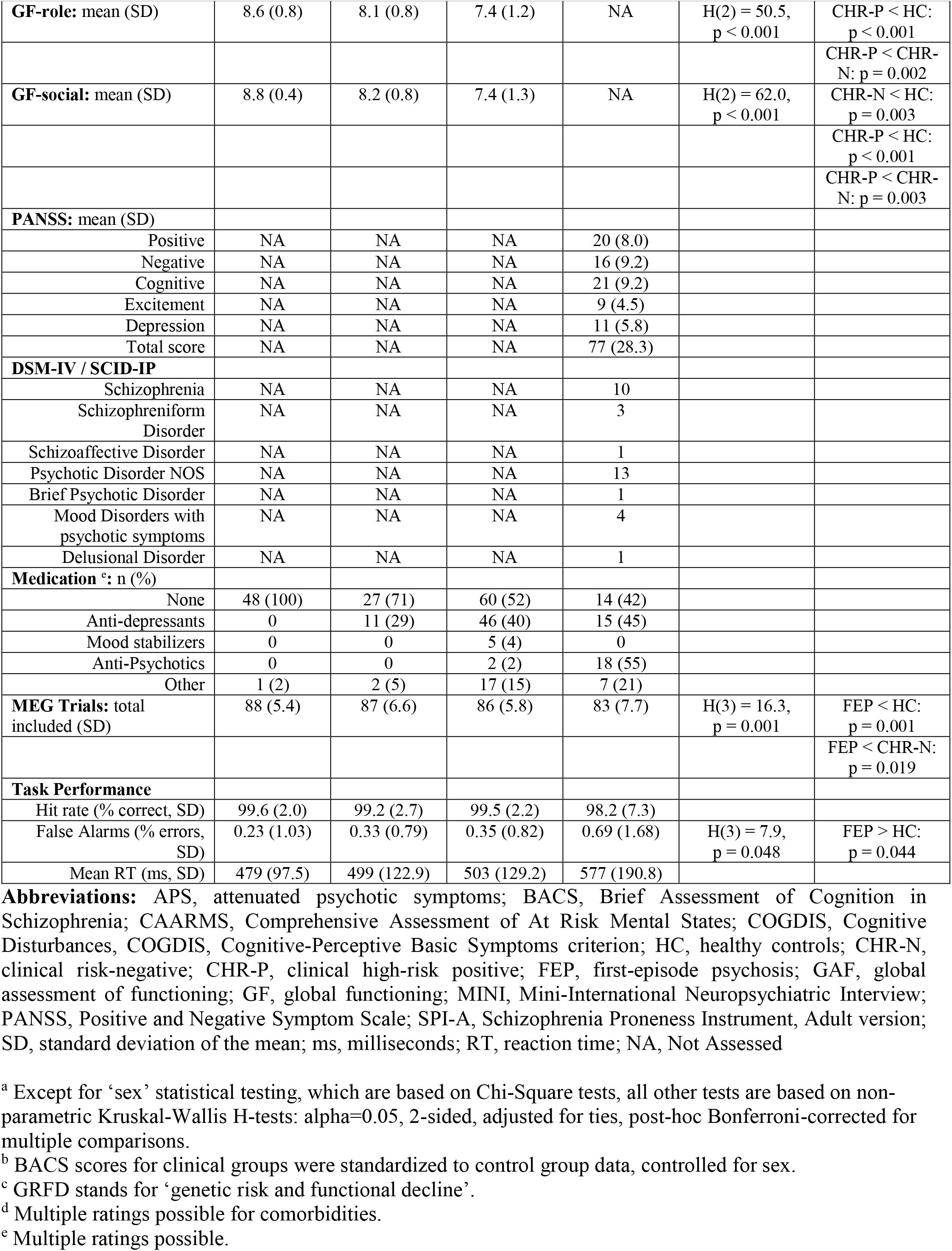
Demographics, Clinical Data, and Task Performance.

The study was approved by the ethical committees of University of Glasgow and the NHS Research Ethical Committee Glasgow & Greater Clyde. All participants provided written informed consent.

### Clinical Follow-Up

Participants meeting CHR-P criteria were re-assessed at 3, 6, 9, 12, 18, 24, 30 and 36 months intervals to examine persistence of Ultra-High-Risk (UHR) criteria and transition to psychosis. Persistence of UHR-criteria was operationalized by the continued presence of APS up to 12 months in this study (see Supplementary materials).

### Stimuli and task

Auditory stimuli were 1000 Hz carrier tones (duration: 2 sec), amplitude modulated (AM) at 40 Hz (23), presented binaurally through inner-ear tubes, with an inter-stimulus-interval (ISI) of on average 2 seconds (jittered between 1.5 and 2.5 seconds, equal distribution). Participants were instructed to fixate a translucent screen (viewing distance: 75 cm). Participants received one block of 100 of 40-Hz AM tones (‘ripple tones’, Figure 1A). To control for potential attentional differences, 10 additional identical sounds with equal intensity levels over time (‘flat tones’) were interspersed, serving as targets to respond to by button-press. These target trials were not included in the analyses of the MEG data.

**Figure 1:**
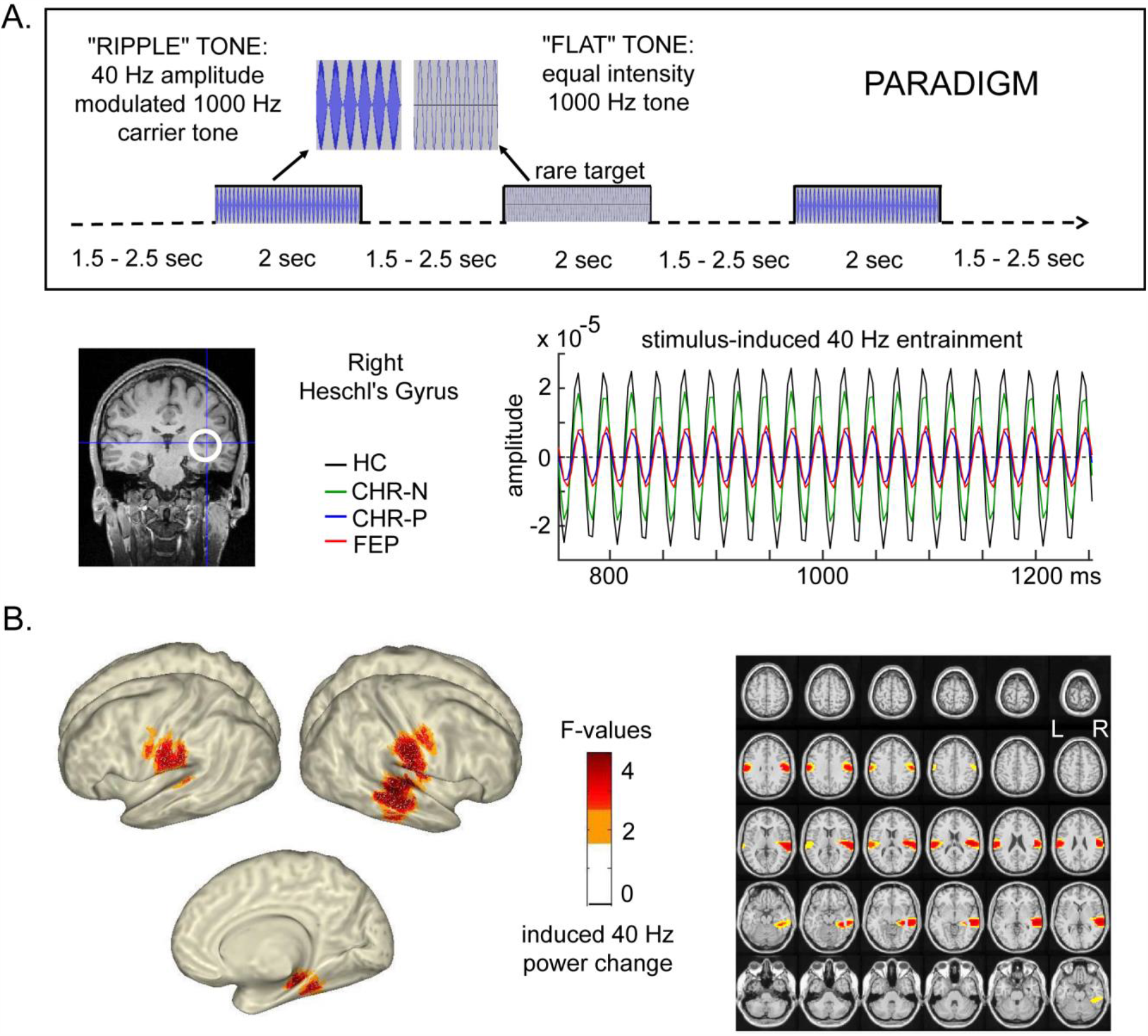
Task paradigm and brain regions showing stimulus-entrained 40-Hz responses. A) Task paradigm and an example of 40-Hz narrow-band-pass filtered signals from the four main groups in right Heschl’s gyrus.Data shows clear entrainment to stimulation frequency in all main groups: HC = healthy controls, CHR-N = clinical-high-risk negative clinical controls, CHR-P = clinical-high-risk positive, FEP = first-episode psychosis patients. B) Whole-brain significant increases of 40-Hz induced power (250-1750 ms versus 500 ms baseline) across all participants. Orange color represents F-values uncorrected for multiple comparisons, whereas the red colors indicate FDR corrected brain activity.

### Neuroimaging

MEG-data were acquired from a 248-channel 4D-BTI magnetometer system (MAGNES® 3600 WH, 4D-Neuroimaging, San Diego), recorded with 1017.25 Hz sampling rate, and DC-400 Hz online filtered. T1 anatomical scans (3D MPRAGE sequences) were collected on a Siemens Trio Tim 3T-scanner (192 slices, voxel size 1 mm^3^, FOV=256×256×176 mm^3^, TR=2250 ms, TE=2.6 ms, FA=9°) for subject-specific source localization of MEG activity.

### Task performance data analysis

Analysis of task data included percentage of correctly detected flat-tone targets, mean reaction times of correct responses and false alarms.

### MEG data analysis

MEG data were analysed with MATLAB 2013b using the open-source Fieldtrip Toolbox Version 20161023 (http://www.fieldtriptoolbox.org/). Epochs of 4 seconds duration (1 second baseline), time-locked to sound onset, were extracted for the task-irrelevant 40-Hz AM tones only (excluding false alarm trials). Line noise contamination was attenuated with a discrete 50-Hz Fourier transform filter. Faulty sensors with large signal variance or flat signals were removed, and data down-sampled to 300 Hz. Artefact-free data were created by removing trials with excessive transient muscle activity, slow drift or SQUID jumps using visual inspection and applying Independent Component Analysis (ICA)-based removal of eye-blink, eye-movement and Electrocardiographic (ECG) artefacts.

Sensor-level data were submitted to time-frequency analyses (frequency resolution 0.25 Hz, 500 ms sliding window, 25 ms step-size, Hanning tapered) using planar-orientation transformed MEG data (31). Corresponding Inter-Trial-Phase-Coherence (ITPC) activity (32) was computed from fourier output. All data were expressed as relative change (relch) from baseline activity (−500-0 ms).

The main analyses, however, focused on data transformed into source-space because regional specificity at each sensor is compromised by field-spread through inputs from multiple sources and inter-individual differences in temporal cortex folding (24). The basic source-level analyses steps included: 1) determination of regions showing stimulus-entrained 40-Hz ASSR signal, 2) determination of a subset of regions showing main group effects (HC, CHR-P, CHR-N, FEP), 3) identification of group differences (post-hoc pairwise group comparisons), and 4) contributions of ASSR impairments towards predicting transition to psychosis and/or 1-year persistence of APS.

First, MEG data were co-registered with the T1 MRI scans, using anatomical landmarks (nasion, bilateral pre-auricular points) and head-shape data collected using a Polhemus 3D Fasttrack digitization system. A three-dimensional grid of 5 mm resolution was then created and linearly warped into a single-shell volume conductor model.

Second, 40-Hz ASSR power (38-42 Hz) between 250-1750 ms, compared to baseline power (−500-0 ms), was source-localized using the Dynamic Imaging of Coherent Sources (DICS) beamforming approach (33), and a common-filters approach. Ten AAL-atlas (34) ROIs (Figure 1B) were then statistically identified as brain regions significantly entrained to the 40-Hz stimulation frequency, including right hemisphere Heschl’s gyrus (RHES), Superior Temporal Gyrus (RSTG), Middle Temporal Gyrus (RMTG), Thalamus (RTHA), Hippocampus (RHIP), and Parahippocampal Gyrus (RPHG), and bilateral Rolandic Operculum (LROL/RROL) and SupraMarginal Gyrus (LSMG/RSMG).

### Statistical analyses

#### Main effects of Stimulation and Group

Main effects of stimulation were determined from DICS whole-brain activity, using Monte-Carlo permutation-based dependent-sample t-tests (alpha=0.05, one-sided, FDR-corrected) across all 236 participants, with baseline and stimulus-induced 40-Hz ASSR power as paired samples. Main effects of group included linear regression analysis (backward-selection method, alpha=0.05, 2-sided, 1000 sample bootstrapped) on 40-Hz amplitude and 40-Hz ITPC virtual-channel data (250-2000 ms averaged relch data) from all regions significantly entrained to the 40-Hz stimulation.

#### Group differences

Group differences in MEG-included trial numbers, behavioral task performance, and demographic and clinical as well as cognition data were assessed with non-parametric Kruskal-Wallis tests, alpha-level 0.05, 2-sided, with post-hoc pairwise comparisons Bonferroni-corrected for Type I errors. BACS data were first z-normalized to the HC data.

For sensor-level ASSR data (power and ITPC relch data), averaged across 39-41 Hz, 250-2000 ms, and 4 right frontal-temporal sensors (see Supplementary materials, Figure S1), Kruskal-Wallis tests were used to test for main group differences.

Virtual-channel 40-Hz ASSR data from entrained brain regions across participants were subjected to a non-parametric permutation (n=5000) t-test, tested against the null hypothesis of no difference from the HC group, using estimation statistics (35). Confidence intervals (CI) were obtained from the central 95% of the resampling distribution. To account for skewed data distribution, data were bias and accelerated bootstrap corrected.

Post-hoc analyses were conducted for CHR-P participants with persistent APS (APS-P) vs. non-persistent symptoms (APS-NP) within 1-year follow-up as well as between CHR-Ps who transitioned to psychosis (CHR-P-T) vs. those who did not (CHR-P-NT). Effect sizes were computed using Cohen’s-d.

#### Correlation and classification analyses

In order to establish predictors for APS-persistence and transition to psychosis, we used a combination of linear regression analysis and linear discriminant analysis. First, linear regression models (backward selection, Confidence Interval [CI] 95%, alpha=0.05, 2-sided, 1000 sample bootstrapping) were computed for the contrast between 1) APS-P (n=40) and APS-NP (n=38), and 2) between CHR-P-T (n=13) and CHR-P-NT (n=103) subgroups, using ITPC and amplitude measures from all ten 40-Hz entrained ROIs as dependent variables. ASSR data were z-normalized to HCs. The resulting set of significant predictors for each of the contrasts were then used in Linear Discriminant Analyses (LDA) to evaluate classification accuracy for APS-persistence and transition to psychosis, based on all significant predictors, and discriminating ability (Standardized Canonical Discriminant Function Coefficients) of separate predictors. Data were cross-validated with a leave-one-out method. For each model, both Wilks’ Lambda/Chi-Square tests as well as area under the Receiver Operating Characteristic Curve (ROC-AUC) were evaluated.

## Results

### Demographic/clinical Data and Task performance

The FEP group included significantly more male participants compared to HC (p=0.004), CHR-N (p=0.002) and CHR-P groups (p<0.001) (Table 1). FEP patients were significantly older than CHR-Ps (p=0.015). CHR-P participants had significantly less years of education (p=0.032) and significantly lower BACS composite (p=0.014), Token Motor (p<0.001) and Symbol Coding (p=0.002) scores compared to HC. FEP, CHR-P and CHR-N all had significantly lower Global Assessment of Functioning (GAF) scores than HC participants and each other (all p-values<0.001). In addition, the CHR-P group had lower scores than HC in global role functioning (p<0.001). Both CHR-P and CHR-N were also characterized by lower social functioning (CHR-N, p=0.003; CHR-P, p<0.001). FEP patients produced more behavioral false alarms than HCs on rare-targets during the ASSR task (p=0.044).

### Follow-Up Outcomes

Forty CHR-Ps continued to meet APS-criteria while 38 did not. Compared to APS-NPs, APS-Ps scored significantly higher on CAARMS-(p=0.001) and SPI-A-severity (p=0.034), as well as significantly lower on global role functioning at baseline (p=0.028) (Table 2).

**Table 2.**
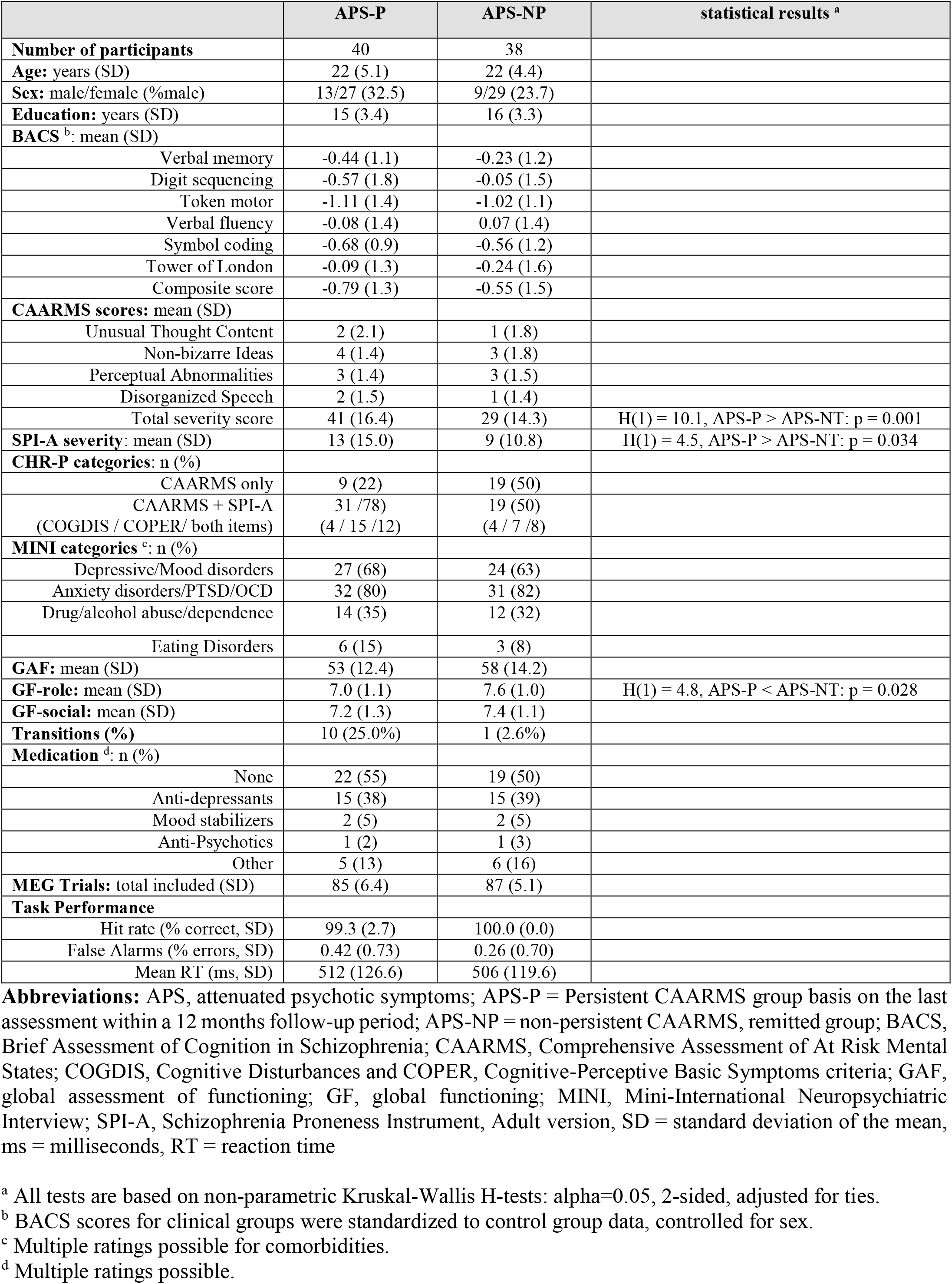
Demographic and Clinical Characteristics for APS-persistent and APS-non-persistent CHR-P participants.

|  | APS-P | APS-NP | statistical results <sup>a</sup> |
| --- | --- | --- | --- |
| **Number of participants** | 40 | 38 |  |
| **Age:** years (SD) | 22 (5.1) | 22 (4.4) |  |
| **Sex:** male/female (% male) | 13/27 (32.5) | 9/29 (23.7) |  |
| **Education:** years (SD) | 15 (3.4) | 16 (3.3) |  |
| **BACS <sup>b</sup>:** mean (SD) |  |  |  |
| Verbal memory | -0.44 (1.1) | -0.23 (1.2) |  |
| Digit sequencing | -0.57 (1.8) | -0.05 (1.5) |  |
| Token motor | -1.11 (1.4) | -1.02 (1.1) |  |
| Verbal fluency | -0.08 (1.4) | 0.07 (1.4) |  |
| Symbol coding | -0.68 (0.9) | -0.56 (1.2) |  |
| Tower of London | -0.09 (1.3) | -0.24 (1.6) |  |
| Composite score | -0.79 (1.3) | -0.55 (1.5) |  |
| **CAARMS scores:** mean (SD) |  |  |  |
| Unusual Thought Content | 2 (2.1) | 1 (1.8) |  |
| Non-bizarre Ideas | 4 (1.4) | 3 (1.8) |  |
| Perceptual Abnormalities | 3 (1.4) | 3 (1.5) |  |
| Disorganized Speech | 2 (1.5) | 1 (1.4) |  |
| Total severity score | 41 (16.4) | 29 (14.3) | H(1) = 10.1, APS-P > APS-NT: p = 0.001 |
| **SPI-A severity:** mean (SD) | 13 (15.0) | 9 (10.8) | H(1) = 4.5, APS-P > APS-NT: p = 0.034 |
| **CHR-P categories:** n (%) |  |  |  |
| CAARMS only | 9 (22) | 19 (50) |  |
| CAARMS + SPI-A<br>(COGDIS / COPER/ both items) | 31 /78<br>(4 / 15 /12) | 19 (50)<br>(4 / 7 /8) |  |
| **MINI categories <sup>c</sup>:** n (%) |  |  |  |
| Depressive/Mood disorders | 27 (68) | 24 (63) |  |
| Anxiety disorders/PTSD/OCD | 32 (80) | 31 (82) |  |
| Drug/alcohol abuse/dependence | 14 (35) | 12 (32) |  |
| Eating Disorders | 6 (15) | 3 (8) |  |
| **GAF:** mean (SD) | 53 (12.4) | 58 (14.2) |  |
| **GF-role:** mean (SD) | 7.0 (1.1) | 7.6 (1.0) | H(1) = 4.8, APS-P < APS-NT: p = 0.028 |
| **GF-social:** mean (SD) | 7.2 (1.3) | 7.4 (1.1) |  |
| **Transitions (%)** | 10 (25.0%) | 1 (2.6%) |  |
| **Medication <sup>d</sup>:** n (%) |  |  |  |
| None | 22 (55) | 19 (50) |  |
| Anti-depressants | 15 (38) | 15 (39) |  |
| Mood stabilizers | 2 (5) | 2 (5) |  |
| Anti-Psychotics | 1 (2) | 1 (3) |  |
| Other | 5 (13) | 6 (16) |  |
| **MEG Trials:** total included (SD) | 85 (6.4) | 87 (5.1) |  |
| **Task Performance** |  |  |  |
| Hit rate (% correct, SD) | 99.3 (2.7) | 100.0 (0.0) |  |
| False Alarms (% errors, SD) | 0.42 (0.73) | 0.26 (0.70) |  |
| Mean RT (ms, SD) | 512 (126.6) | 506 (119.6) |  |

**Abbreviations:** APS, attenuated psychotic symptoms; APS-P = Persistent CAARMS group basis on the last assessment within a 12 months follow-up period; APS-NP = non-persistent CAARMS, remitted group; BACS, Brief Assessment of Cognition in Schizophrenia; CAARMS, Comprehensive Assessment of At Risk Mental States; COGDIS, Cognitive Disturbances and COPER, Cognitive-Perceptive Basic Symptoms criteria; GAF, global assessment of functioning; GF, global functioning; MINI, Mini-International Neuropsychiatric Interview; SPI-A, Schizophrenia Proneness Instrument, Adult version, SD = standard deviation of the mean, ms = milliseconds, RT = reaction time
<sup>a</sup> All tests are based on non-parametric Kruskal-Wallis H-tests: alpha=0.05, 2-sided, adjusted for ties.
<sup>b</sup> BACS scores for clinical groups were standardized to control group data, controlled for sex.
<sup>c</sup> Multiple ratings possible for comorbidities.
<sup>d</sup> Multiple ratings possible.

Out of 116 CHR-P participants, 13 participants (11.2 %) made a transition to psychosis (mean follow-up period: 18 months [6-36 months], Table 3). Transitioned CHR-Ps had significantly lower GAF (p=0.034) and global social functioning scores (p=0.023) at baseline than the CHR-P-NT group.

**Table 3.**
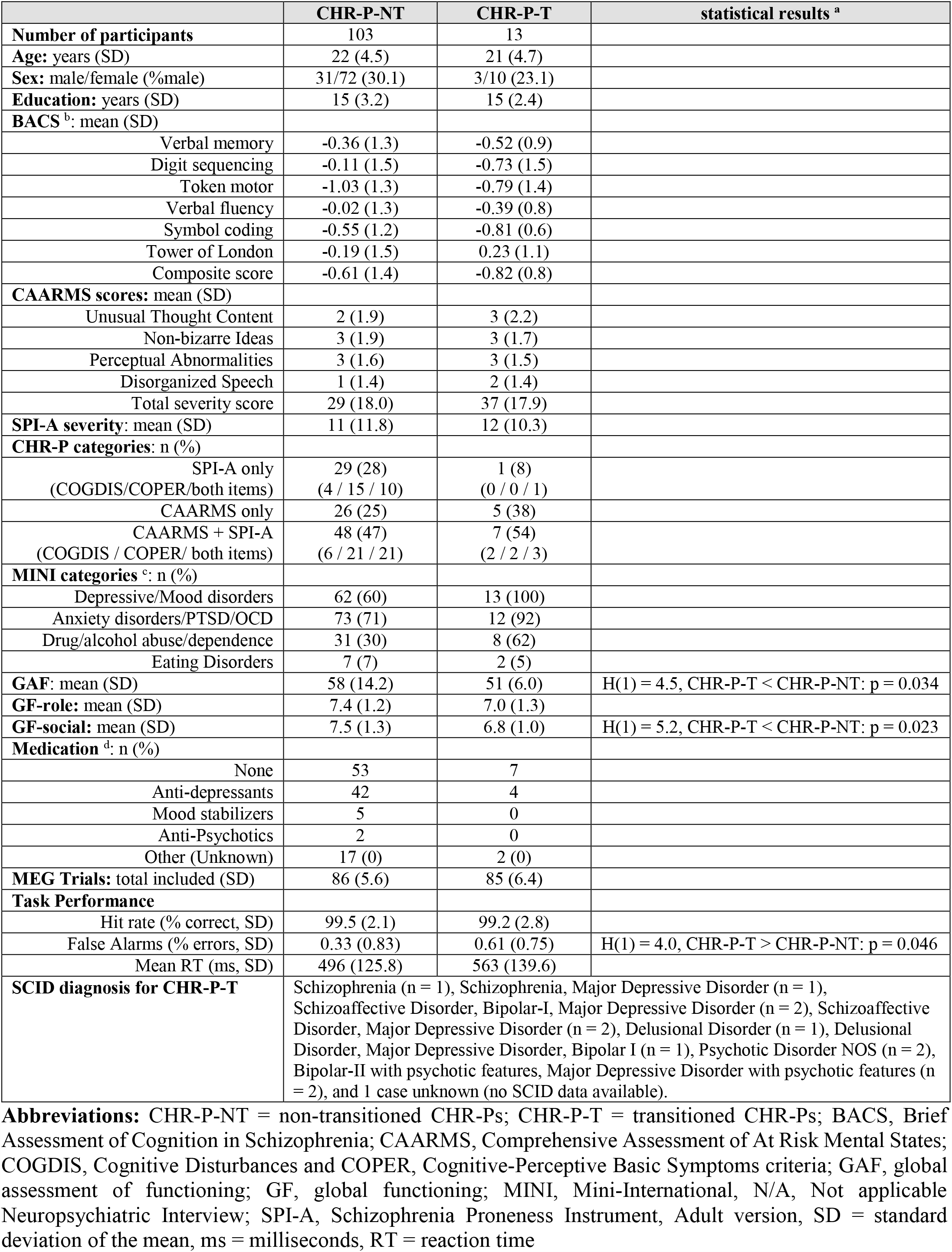
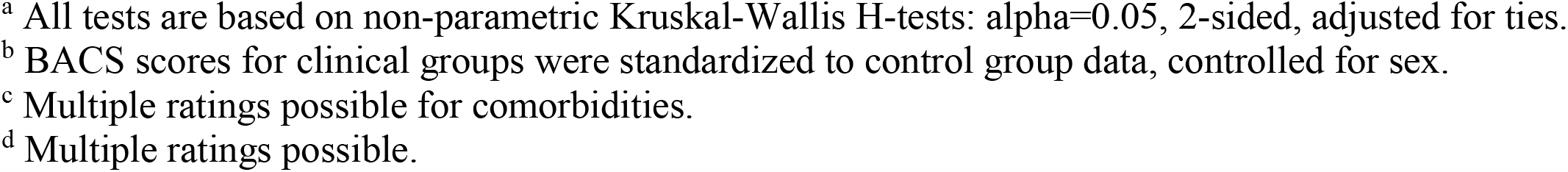
Demographic and Clinical Characteristics for Transitioned and Non-transitioned CHR-P participants.

|  | **CHR-P-NT** | **CHR-P-T** | **statistical results <sup>a</sup>** |
| --- | --- | --- | --- |
| **Number of participants** | 103 | 13 |  |
| **Age:** years (SD) | 22 (4.5) | 21 (4.7) |  |
| **Sex:** male/female (% male) | 31/72 (30.1) | 3/10 (23.1) |  |
| **Education:** years (SD) | 15 (3.2) | 15 (2.4) |  |
| **BACS <sup>b</sup>:** mean (SD) |  |  |  |
| Verbal memory | -0.36 (1.3) | -0.52 (0.9) |  |
| Digit sequencing | -0.11 (1.5) | -0.73 (1.5) |  |
| Token motor | -1.03 (1.3) | -0.79 (1.4) |  |
| Verbal fluency | -0.02 (1.3) | -0.39 (0.8) |  |
| Symbol coding | -0.55 (1.2) | -0.81 (0.6) |  |
| Tower of London | -0.19 (1.5) | 0.23 (1.1) |  |
| Composite score | -0.61 (1.4) | -0.82 (0.8) |  |
| **CAARMS scores:** mean (SD) |  |  |  |
| Unusual Thought Content | 2 (1.9) | 3 (2.2) |  |
| Non-bizarre Ideas | 3 (1.9) | 3 (1.7) |  |
| Perceptual Abnormalities | 3 (1.6) | 3 (1.5) |  |
| Disorganized Speech | 1 (1.4) | 2 (1.4) |  |
| Total severity score | 29 (18.0) | 37 (17.9) |  |
| **SPI-A severity:** mean (SD) | 11 (11.8) | 12 (10.3) |  |
| **CHR-P categories:** n (%) |  |  |  |
| SPI-A only<br>(COGDIS/COPER/both items) | 29 (28)<br>(4 / 15 / 10) | 1 (8)<br>(0 / 0 / 1) |  |
| CAARMS only | 26 (25) | 5 (38) |  |
| CAARMS + SPI-A<br>(COGDIS / COPER/ both items) | 48 (47)<br>(6 / 21 / 21) | 7 (54)<br>(2 / 2 / 3) |  |
| **MINI categories <sup>c</sup>:** n (%) |  |  |  |
| Depressive/Mood disorders | 62 (60) | 13 (100) |  |
| Anxiety disorders/PTSD/OCD | 73 (71) | 12 (92) |  |
| Drug/alcohol abuse/dependence | 31 (30) | 8 (62) |  |
| Eating Disorders | 7 (7) | 2 (5) |  |
| **GAF:** mean (SD) | 58 (14.2) | 51 (6.0) | H(1) = 4.5, CHR-P-T < CHR-P-NT: p = 0.034 |
| **GF-role:** mean (SD) | 7.4 (1.2) | 7.0 (1.3) |  |
| **GF-social:** mean (SD) | 7.5 (1.3) | 6.8 (1.0) | H(1) = 5.2, CHR-P-T < CHR-P-NT: p = 0.023 |
| **Medication <sup>d</sup>:** n (%) |  |  |  |
| None | 53 | 7 |  |
| Anti-depressants | 42 | 4 |  |
| Mood stabilizers | 5 | 0 |  |
| Anti-Psychotics | 2 | 0 |  |
| Other (Unknown) | 17 (0) | 2 (0) |  |
| **MEG Trials:** total included (SD) | 86 (5.6) | 85 (6.4) |  |
| **Task Performance** |  |  |  |
| Hit rate (% correct, SD) | 99.5 (2.1) | 99.2 (2.8) |  |
| False Alarms (% errors, SD) | 0.33 (0.83) | 0.61 (0.75) | H(1) = 4.0, CHR-P-T > CHR-P-NT: p = 0.046 |
| Mean RT (ms, SD) | 496 (125.8) | 563 (139.6) |  |
| **SCID diagnosis for CHR-P-T** | Schizophrenia (n = 1), Schizophrenia, Major Depressive Disorder (n = 1), Schizoaffective Disorder, Bipolar-I, Major Depressive Disorder (n = 2), Schizoaffective Disorder, Major Depressive Disorder (n = 2), Delusional Disorder (n = 1), Delusional Disorder, Major Depressive Disorder, Bipolar I (n = 1), Psychotic Disorder NOS (n = 2), Bipolar-II with psychotic features, Major Depressive Disorder with psychotic features (n = 2), and 1 case unknown (no SCID data available). |  |  |

**Abbreviations:** CHR-P-NT = non-transitioned CHR-Ps; CHR-P-T = transitioned CHR-Ps; BACS, Brief Assessment of Cognition in Schizophrenia; CAARMS, Comprehensive Assessment of At Risk Mental States; COGDIS, Cognitive Disturbances and COPER, Cognitive-Perceptive Basic Symptoms criteria; GAF, global assessment of functioning; GF, global functioning; MINI, Mini-International, N/A, Not applicable Neuropsychiatric Interview; SPI-A, Schizophrenia Proneness Instrument, Adult version, SD = standard deviation of the mean, ms = milliseconds, RT = reaction time
<sup>a</sup> All tests are based on non-parametric Kruskal-Wallis H-tests: $\alpha=0.05$ , 2-sided, adjusted for ties.
<sup>b</sup> BACS scores for clinical groups were standardized to control group data, controlled for sex.
<sup>c</sup> Multiple ratings possible for comorbidities.
<sup>d</sup> Multiple ratings possible.

### Main effect of 40-Hz Stimulation and Group Effects

Across all groups, 40-Hz AM tones induced increased sustained (250-2000 ms) 40-Hz power (Figure 1B, lower panel) in ten brain regions (F-values=2.5-4.6, p=0.002, 95% CI=[-0.042 0.005], FDR corrected), including RHES, RSTG, RMTG, RROL, RSMG, RHIP, RPHG, RTHA, LROL, and LSMG. For these ROIs, virtual channel time-series data were computed using Linearly Constrained Minimum Variance (LCMV) beamformers (36), with normalized lead-fields and a regularisation parameter of 20% to attenuate leakage from nearby sources. Time-series were computed separately for each voxel within each ROI, and then combined into one time-series per ROI using the first Singular Value Decomposition component across the single-voxel data. Data were then submitted to time-frequency fourier analysis (frequency resolution 0.25 Hz, sliding window of 500 ms, step-size 25 ms, Hanning tapered) and ITPC was computed. In addition, 40-Hz ASSR amplitude responses were computed by band-pass filtering single-trial virtual-channel data with a sharp Butterworth filter (range 39.6-40.4 Hz, two-pass, roll-off = 4, see Figure 1A). The filtered data were enveloped and averaged across trials. All virtual-channel data were expressed as relative change (relch) from baseline activity (−500-0 ms).

Linear Regression analyses (backward selection) on 40-Hz amplitude virtual-channel data was used to help select a limited set of ROIs (out of the total 10 stimulus-entrained ROIs) for post-hoc analyses of group differences. For the 40-Hz ITPC data, a significant model was found including RHES activity only (F(1,234)=5.15, p=0.024, adj. R-square=0.017). For 40-Hz amplitude data, a combination of all but the LROL ROI data contributed significant variance to explaining main group differences (F(8,235)=2.17, p=0.031, adj. R-square=0.045). As including all ROIs would increase Type I errors, we decided to further restrict the post-hoc analysis to a subset of ROIs that together explained the main group effect at a Bonferroni corrected alpha-level of 0.0055 (0.05/9). Such a model was found for amplitude changes in RHES, RSTG, RTHA and RHIP (F(4,231)=3.95, p=0.004, adj. R-square=0.048).

### Main Group differences at sensor-level

Sensor-level data showed stimulus-induced 40-Hz power and ITPC increases over right and left frontal-temporal sensors (Figure S1). Compared to HC group, both power and ITPC in the 40-Hz range were reduced over right temporal sensors (Figure S1, bottom three rows) in CHR-P and FEP, but not in the CHR-N group. However, these effects were not significant (Supplementary Materials).

### Main Group differences at source-level

At source-level, post-hoc estimation statistics (Figure 2A/B) of 40-Hz amplitude data from RHES, RSTG, RTHA, and RHIP as well as 40-Hz ITPC data from RHES were used to evaluate differences between clinical groups (CHR-N, CHR-P, and FEP) and HC. Compared to HC, 40-Hz amplitude in RTHA and RHIP was significantly reduced in both FEP and CHR-P participants (RTHA-FEP: −3.71, 95% CI: −6.76 to −0.82, p=0.028, d=0.52; RHIP-FEP: −3.27, 95% CI: −6.14 to −0.39, p=0.037, d=0.49; RTHA-CHR-P: −3.28, 95% CI: −6.20 to −1.2, p=0.003, d=0.47; RHIP-CHR-P: −2.45, 95% CI: −4.98 to −0.32, p=0.019, d=0.37). In addition, the FEP group was impaired in RHES amplitude (−3.31, 95% CI: −6.32 to −0.37, p=0.044, d=0.48) while CHR-P participants were impaired in RHES ITPC (−0.45, 95% CI: −0.95 to −0.04, p=0.043, d=0.34). No impairments were found for CHR-N participants.

**Figure 2.**
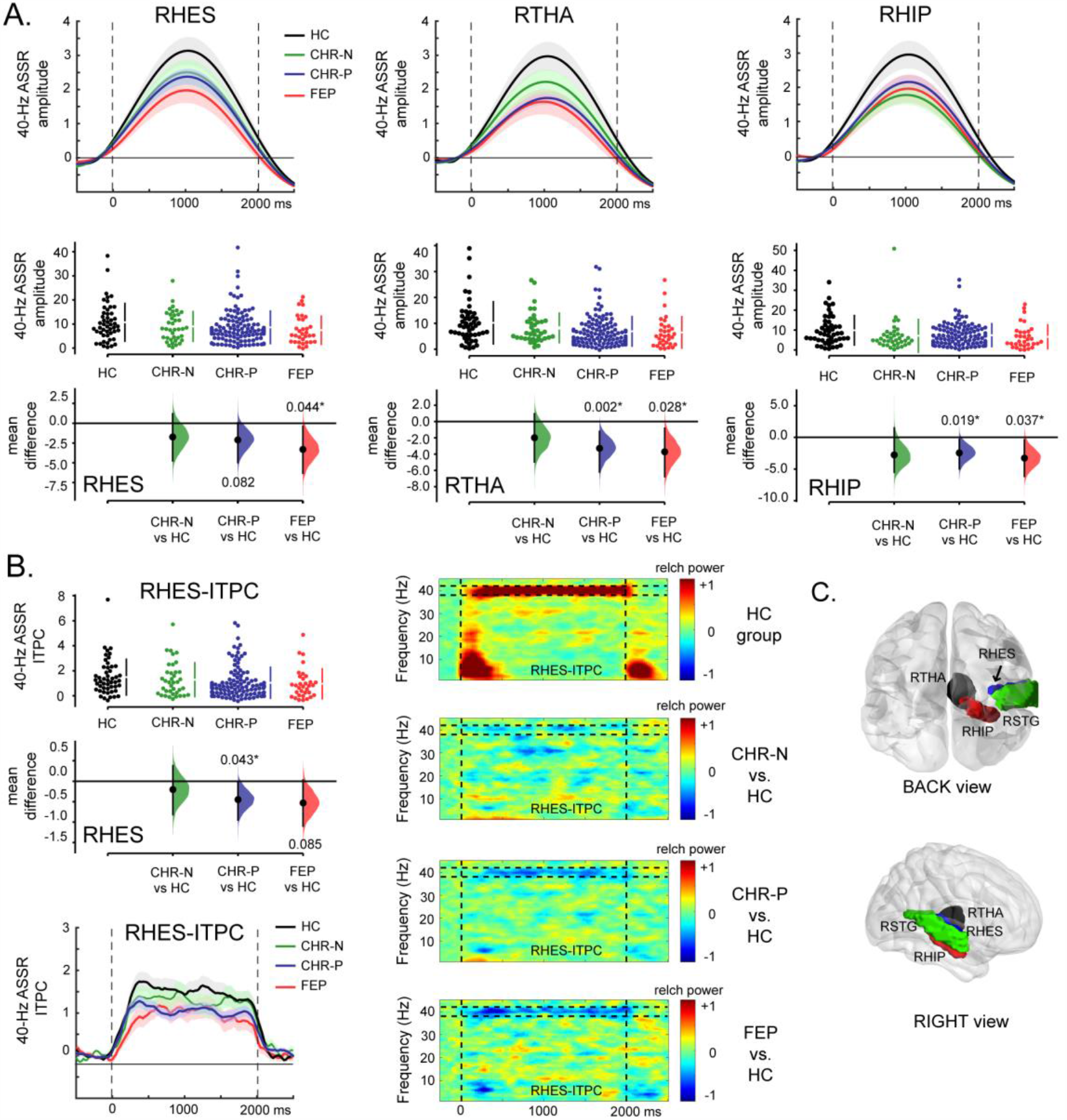
Main group differences in the 40-Hz ASSR signals. A) Top row are group averaged 40-Hz amplitude data (relative-change from baseline) from RHES, RTHA and RHIP, with error bars representing standard-error-of-the-mean (SEM). Stimulus onset at time 0 and offset at 2000 ms. Below the respective Cumming Estimation Plots with data distribution swarm plots and group difference data distributions (compared to healthy controls). Significant contrasts are indicated by an asterisk. B) As panel A but for ITPC data in RHES. In addition, Time-Frequency plots of ITPC (Inter-Trial-Phase-Coherence) data are shown for RHES for HC, and the contrasts of HC with all three main clinical groups. Data represents relative change from baseline, similar to the amplitude data. C) The main 4 ROIs for which virtual channel data were computed and statistically examined for group differences. Abbrev.: HC = healthy controls, CHR-N = clinical-high-risk negative, CHR-P = clinical-high-risk positive, FEP = First-Episode Psychosis, RHES = Right Heschl’s gyrus, RSTG = Right Superior Temporal gyrus, RTHA = Right Thalamus, and RHIP = Right Hippocampus.

### 40-Hz ASSR and follow-up outcomes

Compared to HC, the persistent APS-P group (Figure 3) was impaired in 40-Hz amplitude in RTHA (−4.27, 95% CI: −7.30 to −1.79, p=0.003, d=0.63), and RHIP (−4.14, 95% CI: −6.68 to −2.01, p=0.001, d=0.71), whereas the non-persistent APS-NP group was impaired only in RHES-ITPC (−0.67, 95% CI: −1.20 to −0.20, p=0.013, d=0.55). Similar to the APS-P group, pronounced impairments in RTHA and RHIP were found also for the transitioned CHR-P-T group (RTHA: −6.25, 95% CI: −8.84 to −3.47, p=0.008, d=1.02; RHIP: −5.35, 95% CI: −7.83 to −2.38, p=0.011, d=0.88), with additional impairments in RHES-ITPC signal (−0.84, 95% CI: −1.32 to −0.29, p=0.039, d=0.76).

**Figure 3.**
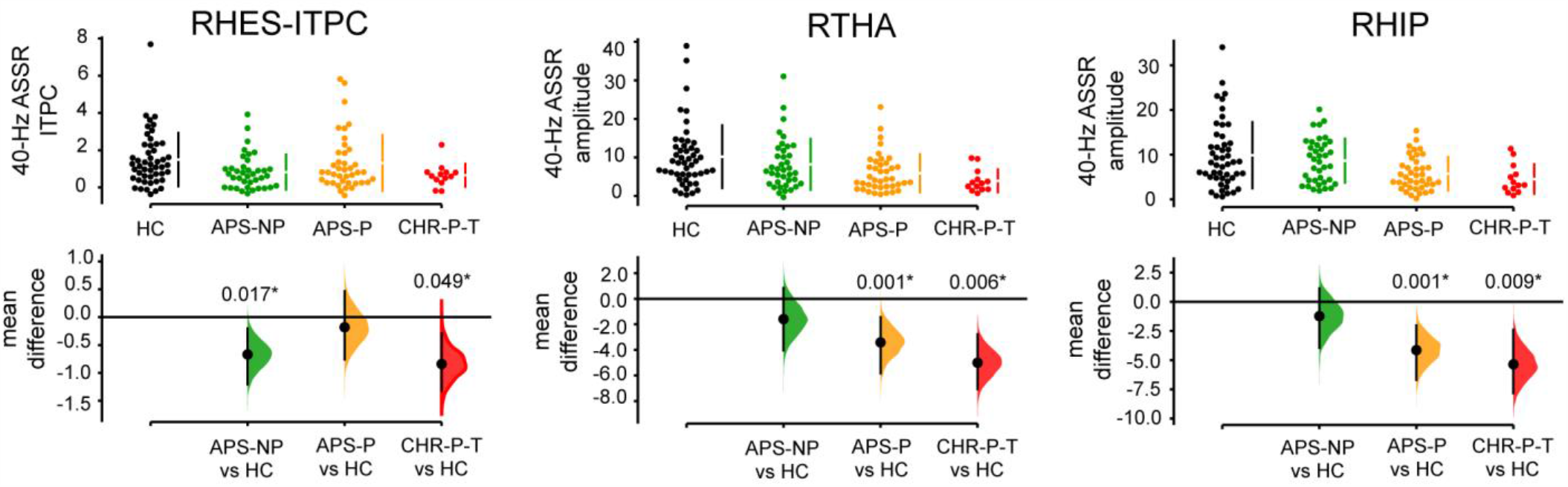
40-Hz ASSR Amplitude Impairments in CHR-P Subgroups. Cumming Estimation Plots of RHES-ITPC, RTHA/RHIP ASSR-amplitude data in follow-up groups, with either presence (APS-P; in green) or absence of persistence of attenuated psychotic symptoms (APS-NP; in orange) up to 1 year follow-up assessment, or based on having transitioned to psychosis (CHR-P-T; in red), with indicated significance (*) from healthy controls (HC). Abbrev.: CHR-P = clinical-high-risk positive participants, APS = attenuated psychotic symptoms, ITPC = Inter-Trial-Phase-Coherence, relch = relative change from baseline (−500-0 ms).

Furthermore, the APS-P group significantly differed from the APS-NP group in RHIP (−2.91, 95% CI: −4.89 to −0.97, p=0.005, d=0.66), but not in RHES (p=0.98) or RTHA (p=0.09). Compared to CHR-P-NT, CHR-P-Ts were significantly impaired both in RTHA (−3.34, 95% CI: −5.03 to −1.04, p=0.043, d=0.72), and in RHIP (−3.27, 95% CI: −5.10 to −0.89, p=0.048, d=0.68), but not in RHES (p=0.39).

As the inclusion of CHR-P-T participants in APS-P (n=10) group and APS-NP group (n=1) could potentially drive the effects reported above for the APS subgroups, we repeated the analyses excluding CHR-P-T data. The APS-P group (n=30) continued to show impairments in RTHA (−3.71, 95% CI: −6.70 to −0.83, p=0.003, d=0.54), and RHIP (−3.90, 95% CI: −6.45 to −1.57, p=0.008, d=0.66), while the non-persistent APS-NP (n=37) group was impaired in RHES-ITPC (−0.66, 95% CI: −1.18 to −0.20, p=0.012, d=0.54), compared to HCs. Finally, we investigated potential effects of anti-psychotic medication (AP) on 40 Hz ASSR amplitude in FEP patients (non-AP-medicated FEPs: n=15; AP-medicated FEPs: n=18). No significant differences were found between groups (RHES: p=0.77; RTHA p=0.64 and RHIP p=0.28).

### Classification of APS-Persistence and Transition in the CHR-P Group

A combination of linear regression (backward method) and LDA was used to find a subset of 40-Hz ASSR measures for classifying and predicting clinical outcomes, using all available amplitude and ITPC data as input data, z-normalized to the HC-data.

Linear regression analyses of persistent (n=40) and non-persistent APS (n=38) groups revealed a significant model (F(5,77)=7.3, p<0.001) including RHIP-amplitude (Beta=−0.338, t=−3.1, p=0.002, 95% CI=[-0.44 − 0.10]), RMTG-amplitude (Beta=−0.225, t=−2.0, p=0.048, 95% CI=[−0.23 −0.01]), RSMG-amplitude (Beta=0.254, t=2.5, p=0.015, 95% CI=[0.03 0.22]), LSMG-ITPC (Beta=0.169, t=1.7, p=0.1, 95% CI=[-0.02 0.22]), and RSTG-ITPC (Beta=0.313, t=2.9, p=0.005, 95% CI=[0.06 0.34]). Including these five predictors revealed a significant classification model (Wilks’ Lambda=0.664, χ(5)=30.1, p<0.001, AUC=0.842) with standardized Canonical Discriminant Function Coefficients (CDFCs) of −0.678 for RHIP-amplitude, −0.469 for RMTG-amplitude, 0.524 for RSMG-amplitude, 0.348 for LSMG-ITPC and 0.636 for RSTG-ITPC. A total of 28 out of 38 APS-NP (73.7%) and 29 out of 40 APS-P (72.5%) of participants were correctly classified. For the 103 non-transitioned (CHR-P-NT) and 13 transitioned (CHR-P-T) participants, a significant linear regression model was found (F(5,115)=3.6, p=0.005), including RHES-amplitude (Beta=0.267, t=2.6, p=0.01, 95% CI=[0.02 0.17]), LROL-amplitude (Beta=0.226, t=2.0, p=0.046, 95% CI=[0.002 0.20]), LROL-ITPC (Beta=−0.251, t=−2.5, p=0.029, 95% CI=[-0.14 −0.01]), RTHA-amplitude (Beta=−0.204, t=−2.0, p=0.045, 95% CI=[-0.18 −0.002]), and RHIP-amplitude (Beta=−0.206, t=−2.2, p=0.036, 95% CI=[-0.16 −0.005]). LDA-analysis including these predictors revealed a significant classification model (Wilks’ Lambda=0.860, χ(5)=16.8, p=0.005, AUC=0.810), with standardized CDFCs of −0.768 for RHES-amplitude, −0.649 for LROL-amplitude, 0.721 for LROL-ITPC, 0.577 for RTHA-amplitude and 0.584 for RHIP-amplitude. A total of 79 out of 103 CHR-P-NT (76.7%) and 7 out of 13 CHR-P-T (53.8%) of participants were correctly classified.

### MEG activity in subcortical regions

Estimation of subcortical activity potentially suffers from source-leakage from cortical sources and/or effects of depth-bias corrections. In order to validate the RTHA and RHIP results, we therefore examined group differences also in LTHA and LHIP. There were no main group differences in these ROIs (LTHA: CHR-N, p=0.31, CHR-P, p=0.41, FEP, p=0.08; LHIP: CHR-N, p=0.06, CHR-P, p=0.73, FEP, p=0.45).

## Discussion

Robust evidence exists for impairments in 40-Hz ASSRs in ScZ-patients (14) which is consistent with evidence for alterations in neural circuits that are involved in the generation of gamma-band rhythms, such as PV+ interneurons and NMDA-R mediated neurotransmission (13, 37). However, it is currently unclear whether 40-Hz ASSRs are impaired during emerging psychosis and can predict clinical outcomes. To address these fundamental issues, we implemented a state-of-the-art MEG-approach to examine which brain regions contribute towards 40-Hz ASSR deficits in a large CHR-P sample and to determine whether such impairments predict persistence of APS-status and transition to psychosis.

Consistent with our hypothesis, we found overlapping reductions in 40-Hz ASSRs in FEP and CHR-P participants in RHES, RTHA and RHIP, three brain regions significantly entrained by our 40-Hz stimulation. Deficits in RHES are in line with previous observations in ScZ that have localized 40-Hz ASSR impairments to primary auditory cortex and superior temporal cortex (38, 39). These results contrast, however, with recent EEG-studies that found intact 40-Hz ASSRs in both CHR-P (18, 19) and FEP-groups (18, 40).

One reason for our different findings may constitute the larger sample size in the current study. We also implemented a novel analysis approach that involved narrow-band-pass filtering of single-trial MEG-activity which, compared to more typically used time-frequency analyses, is less affected by trade-offs in time/frequency-resolution. Importantly, the use of MEG allowed us to compute virtual-channel source-level ASSRs from cortical and subcortical regions entrained to the stimulation frequency, thus increasing SNR of ASSR-estimates (23).

Interestingly, our whole-brain source-level approach revealed impairments beyond cortical auditory processing areas in CHR-Ps and FEP-groups, in right thalamus and right hippocampus. The significant thalamic contribution towards 40-Hz ASSRs deficits is consistent with several lines of evidence. Thalamic 40-Hz ASSRs have been observed with PET, fMRI and EEG/MEG (8, 9, 41). Furthermore, experimentally induced thalamic evoked gamma oscillations have been shown to have a direct effect on auditory cortex ASSR responses to click trains (42). Similarly, the hippocampus exhibits an intrinsic 40-Hz rhythm that can be entrained by both internal network (43) as well as external signals such as ASSR stimulation (7, 44).

Importantly, there is consistent evidence for circuit changes implicating deficits in GABAergic neurotransmission in ScZ in auditory as well as thalamic and hippocampal regions. PV+ Interneurons are reduced in hippocampus (45) and thalamic reticular nucleus (46) in ScZ, while in auditory cortex levels of the GABA-synthesizing enzyme GAD65 are decreased (47). Altered synaptic proteins implicating α-amino-3-hydroxy-5-methyl-4-isoxazolepropionic acid (AMPA) receptor-subunits but not NMDA-Rs have been shown to be altered in auditory cortex in ScZ (48). Currently, the precise contributions of Glutamatergic neurotransmission and GABAergic interneurons towards circuit deficits and aberrant gamma-band oscillations remains unclear, however. One possibility is that circuit deficits are due to a primary dysfunction in inhibitory interneurons in ScZ (49). In addition, evidence exists that impaired inhibition could be the result of NMDA-R hypo-functioning on PV+ interneurons (50) or reduced NMDA-R drive on pyramidal cells (51). Crucially, our findings that ASSR-impairments in CHR-Ps represents a potential biomarker for predicting clinical outcomes. Specifically, we show that reductions in 40-Hz ASSRs predicted transition to psychosis in CHR-Ps with good accuracy similar to data from ERPs (52) or MRI (53). In addition, MEG-recorded 40-Hz ASSRs dissociated CHR-Ps with persistence of APS-symptoms at 12-months from those who remitted with excellent accuracy (AUC of 0.842). The finding that ASSR amplitude predicts persistence of APS is an important finding, since only a minority of CHR-P participants will develop psychosis (54) and a large number of individuals will remit from CHR-P status (55). Finally, we did not find 40-Hz ASSR impairments in CHR-N individuals, suggesting that impaired gamma-band oscillations are specifically associated with the CHR-P and FEP phenotypes.

### Strengths and Limitations

There are several limitations of the current study. Firstly, we localized MEG-activity to the thalamus and hippocampus. While localization of subcortical generators remains challenging, we would like to note that reconstruction of MEG time-courses in the thalamus and hippocampus have been demonstrated before by our (56, 57) and others (58, 59). Moreover, our analyses revealed that there were no differences in similar depth regions in the opposite hemisphere, suggesting that for example signal-leakage did not contribute to our observations. Secondly, the sample size of transitioned cases was relatively small (n=13) and would thus require replication in a larger sample.

In conclusion, the current study provides novel evidence on deficits in 40-Hz ASSRs in CHR-P participants and FEP-patients in auditory as well as thalamic and hippocampal regions. Crucially, the current findings highlight that 40-Hz ASSRs predict clinical outcomes in CHR-P participants, including transition to psychosis as well as persistence of APS. Together these findings highlight the potential of MEG as a novel approach to identify circuit dysfunctions and biomarker for clinical outcomes in psychosis.

## Data Availability

Data will be made available upon reasonable request.

## Disclosures and acknowledgments

Dr. Uhlhaas has received research support from Lilly and Lundbeck outside the submitted work. Dr. Lawrie has received a personal fee from Sunovion outside the submitted work. The study was supported by the Medical Research Council (MR/L011689/1). Dr. Rajeev Krishnadas was supported by the Neurosciences Foundation.

We thank Frances Crabbe for help in the acquisition of MEG/MRI-data. The investigators also acknowledge the support of the Scottish Mental Health Research Network (http://www.smhrn.org.uk) now called the NHS Research Scotland Mental Health Network (NRS MHN: http://www.nhsresearchscotland.org.uk/research-areas/mental-health) for providing assistance with participant recruitment, interviews, and cognitive assessments. We would like to thank both the participants and patients who took part in the study and the research assistants of the YouR-study for supporting the recruitment and assessment of CHR-participants.

