## Supplementary Method materials and Figure S1 for "40 Hz Auditory Steady-State Responses Predict Clinical Outcomes in Clinical-High-Risk Participants: A MEG Study"

**Follow-Up assessments**

Participants meeting CHR-P criteria were re-assessed at 3, 6, 9, 12, 18, 24, 30 and 36 months intervals to examine persistence of Ultra-High-Risk (UHR) criteria and transition to psychosis. Persistence of UHR-criteria was operationalized as the continued presence of APS up to 12 months. For 78 CHR-Ps, at least one follow-up assessment within a one-year period was available (n = 60 at 12 month). Criteria for transition to psychosis were defined on the basis of the CAARMS symptom scores of sufficient duration and frequency (symptoms minimally 1 hour/day, 3-6 times/week, > 1 week present), using symptom severity (5-6 out of 6) and frequency scores. When transition to psychosis was confirmed, a SCID Interview was conducted to establish the DSM-IV-category of the psychotic disorder.

**SENSOR-LEVEL analysis of 40 Hz-ASSR data**

Our main analyses focused on data transformed into source space because regional specificity at each sensor is compromised by field-spread through inputs from multiple sources. Also, inter-individual differences in temporal cortex folding tend to create large variance in source projections to the scalp of auditory-cortex activity, which diminishes sensitivity to find group differences at sensor-level. Sensor-level TFR and ITPC data, however, was analysed and results are presented below.

Figure S1 shows the main power (left column) and ITPC responses (right column) for the HC group (top row), with data averaged across a subset of four sensors over right frontal-temporal sites (white dots in top middle topographical distribution plot of planar-transformed data). The three lower rows of the figure show the data for the main group contrasts between clinical groups and controls.

**Figure S1: Main group differences in the 40-Hz ASSR signals at sensor level.**

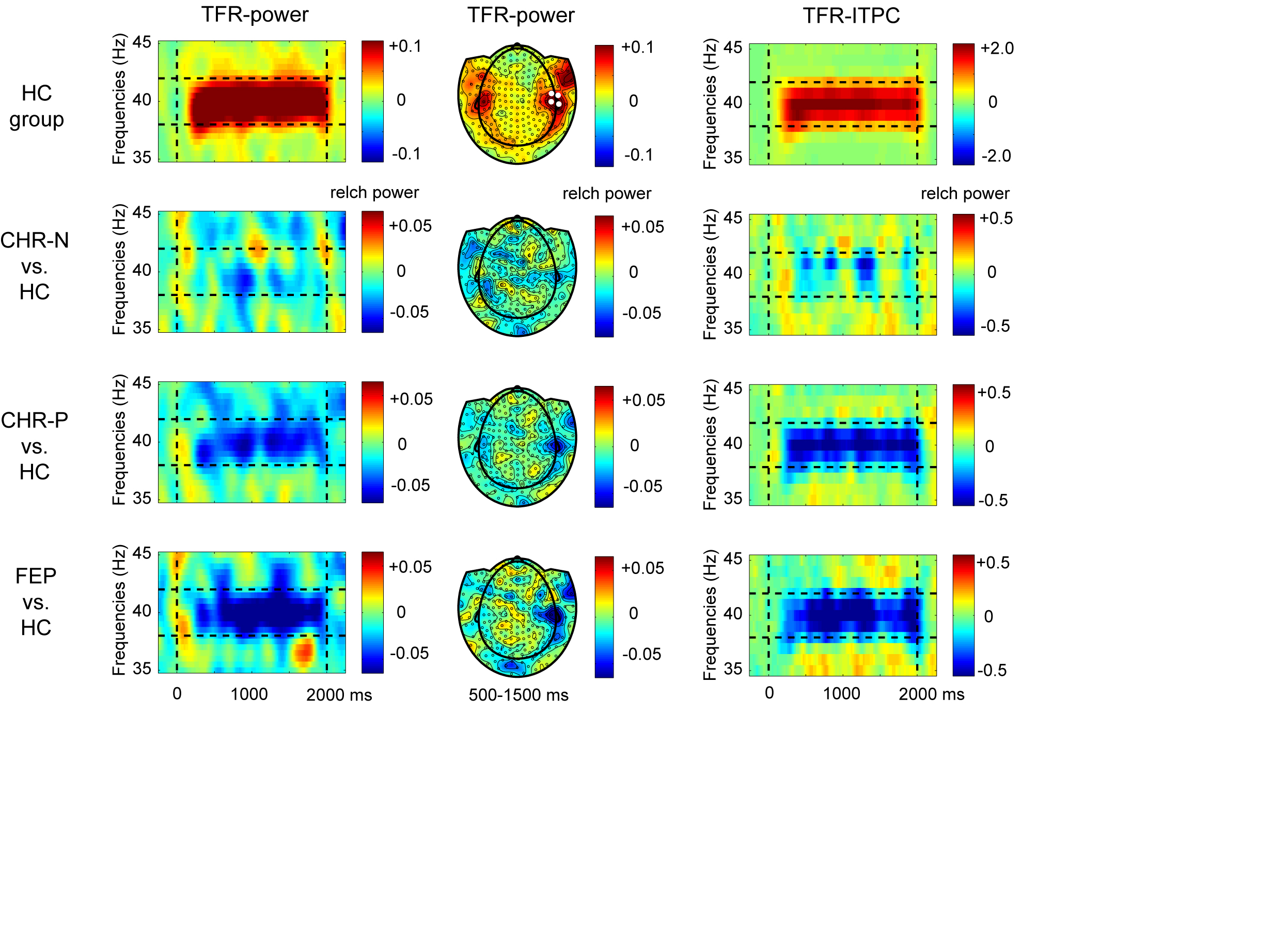

Abbrev.: HC = healthy controls, CHR-N = clinical-high-risk negative, CHR-P = clinical-high-risk positive, FEP = First-Episode Psychosis, relch = relative change from baseline (-500-0 ms) activity

A reduction in 40 Hz-power and 40 Hz-ITPC can be seen over right frontal-central sensors for CHR-P and FEP groups, but not for the CHR-N group. Non-parametric Kruskal Wallis tests were used on data averaged over frequency (39-41 Hz), time (250-2000 ms) and 4 sensors over right auditory regions (indicated in the middle top-row distribution plot of HC group data), to test for main group differences. There were no significant group effects in either TFR (H(3) = 3.9, p = 0.27) or ITPC data H(3) = 4.8, p = 0.18.
